# Chronic Kidney Disease in Jamaica: Estimated Prevalence and Associated Risk Factors from the Jamaica Health and Lifestyle Survey III

**DOI:** 10.1101/2025.03.31.25324786

**Authors:** Lori-Ann M Fisher, Trevor S Ferguson, Kerne D Rocke, Natalie G Guthrie-Dixon, Novie OM Younger-Coleman, Marshall K Tulloch-Reid, Shelly-Ann R McFarlane, Damiann K Francis, Nadia R Bennett, Colette A Cunningham-Myrie, Isthar O Govia, Donovan A McGrowder, William D Aiken, Andriene Grant, Tamu Davidson, Karen Webster-Kerr, Rainford J Wilks

## Abstract

**Introduction:** Jamaica has a high attributable burden of chronic kidney disease (CKD) but no population-based prevalence estimates. We aimed to estimate the prevalence of CKD and explore associated factors.

**Methods:** A secondary analysis of data from Jamaican residents aged ≥15 years from the nationally representative Jamaica Health and Lifestyle Survey-III was performed. CKD was defined as an estimated glomerular filtration rate (eGFR) <60mL/min/1.73 m^2^, using the CKD Epidemiology Collaboration (CKD-EPI) 2021 or Schwartz-Lyon equations, and/or albuminuria ≥30 mg/g. Associated factors included age, sex, socio-economic status, education level, body mass index, hypertension, diabetes mellitus, and sickle cell trait. Weighted prevalence estimates were determined accounting for survey design. Multivariable logistic regression was used to evaluate CKD associations.

**Results:** Analyses included 583 participants, 217 males, mean ±SD age was 49.0 ± 18.2 years. CKD prevalence was 14.8% [95%CI: 11.5%-18.9%]. Seven percent (7.2% [95%CI: 5.1%-10.1%]) had CKD Stage 3 or higher and 8.8% [95%CI:6.3%-12.0%] had albuminuria. CKD participants were older (mean age 57 versus 46.3 years, p<0.001), had higher mean systolic blood pressure (140.3 mmHg versus 128.3 mmHg, p<0.001), and fasting glucose (6.7 micromoles/L versus 5.8 micromoles/L, p<0.001). In a multivariable regression model, hypertension (OR 2.14, 95%CI: 1.22-3.75), diabetes mellitus (OR 2.39, 95%CI: 1.36-4.19) were associated with CKD. Higher education level was inversely associated with CKD, (OR 0.47, 95%CI:0.25-0.89) and (OR 0.41, 95CI: 0.18-0.96) for secondary and tertiary education respectively.

**Conclusion:** An estimated 1 in 7 Jamaicans have CKD. This may translate to increased health care burden on the Jamaican health system.

## Introduction

Chronic kidney disease (CKD) is an emerging global health problem and affects approximately one in ten persons worldwide, resulting in over 1.2 million deaths annually(1, 2). Increased urinary albumin excretion (≥30 mg per day) or reduced glomerular filtration rate (<60mL/min/1.73m^2^) that lasts 3 or more months clinically defines CKD (3). End Stage Kidney failure (ESKF) may develop along the life course of CKD, in which dialysis or transplantation is needed for survival(3). Excess cardiovascular mortality, health care costs, cognitive impairment and poor quality of life are associated with CKD, these risks increasing as estimated glomerular filtration rate (eGFR) declines and albuminuria increases(4–8).

Two thirds of the global CKD population reside in middle and low income countries(9), with Latin America and the Caribbean amongst the highest mortality and disability-adjusted life years attributable to CKD worldwide(1,10), yet sparse population based prevalent data. Jamaica, a middle income Caribbean country, Gross domestic product US 8960 per capita,(10) shares the high burden of CKD described regionally(10, 11). CKD is the fourth leading cause of combined death and disability, representing a 20% increase in the last decade(12). Estimates from the Global Burden of Diseases, CKD Study (1), and the Global Kidney Health Atlas (13), indicate a 9-10% prevalence of CKD in Jamaica(1, 13).However these estimates are derived from complex models based on geographic proximity from countries with robust CKD registry data or surveillance systems or modelled from systematic literature review of available data(1).

CKD risk factors are increasing in Jamaica, one in three Jamaicans have hypertension, 12% with diabetes mellitus and over 50% are overweight or obese(14). Furthermore higher rates of systemic lupus erythematosus, sickle cell disease and trait confer additional risk(11, 15). Prior reports of CKD epidemiology are based on data from the Caribbean renal registry which are likely underestimates due to both the reliance on voluntary reporting and the variability in CKD definitions(11, 16, 17). Ferguson et al in a cross-sectional analysis from a single centre study of 132 diabetic patients attending a sub-speciality clinic, twenty-two percent (22%) had reduced eGFR, and 80%, albuminuria(18). From a cross sectional analysis from the Jamaica Birth Cohort an estimated 8% of persons aged 18-20 years had either albuminuria or reduced eGFR (19). This may suggest a higher than prior reported rate of CKD and highlights the need for population based estimates of CKD prevalence. We aimed to estimate the prevalence of CKD in Jamaica and investigate the association with demographic, clinical and socioeconomic factors in a nationally representative sample.

## Materials and Methods

A cross-sectional secondary analysis of data from the Jamaica Health and Lifestyle Survey 2016-2017 (JHLS-III,), was performed. JHLS-III was a community-based national survey of non-institutionalised persons, resident in Jamaica, aged 15 and older, conducted from 2016 to 2017. The details of the JHLS-III study protocol are published elsewhere(14, 20, 21). The multi-stage sample resulted from randomly selected enumeration districts (EDs) stratified by parish, systematic sampling of twenty households from each ED and selection of a household participant within EDs was done using the Kish Method(24). Ethical approval was obtained from the Ministry of Health and Wellness (MOHW) of Jamaica and The University of the West Indies (UWI) Mona Campus Research Ethics Committees, Ethics Approval number (UWI - ECP 25, 15/16; MOHW-2015/51). All participants provided written informed consent.

Data collectors were trained and certified in questionnaire administration and measurement techniques. Demographics, self-reported medical and medication history, education level and smoking status were collected from questionnaire. Point-of-care blood samples for fasting blood glucose, cholesterol and glycosylated haemoglobin levels; early morning urine samples for albuminuria testing; blood pressure (BP) measurements; weight and height were measured. Venous sampling for serum creatinine was performed. A total of 2807 Jamaicans completed the questionnaire of which 1189 had data for ED, height, age, creatinine and sex and survey weighting for prevalence estimates. Six-hundred and ninety had albuminuria testing. The final sample included in the analyses comprised 583 participants.

## Measurements

Blood pressure was measured using an oscillometric device (Omron 5 Series Blood Pressure Monitor). Three measurements were taken in the right arm after the participant had been seated for 5 minutes, and followed standardised procedures developed for the International Collaborative Study of Hypertension in Blacks(23).

Weight was measured using a portable digital scale (Tanita HD-351 Digital Weight Scale). Height was measured using a portable stadiometer (Seca 213). Fasting glucose and total cholesterol were measured from a capillary blood sample using a point of care device (SD LipidoCare).

Serum creatinine from a single blood sample was measured using a Cobas c111 analyser and urine creatinine from an early morning urinary sample using a Cobas 6000 analyser at the Tropical Metabolism Research Unit (TMRU) Laboratory in the Caribbean Institute for Health Research, Mona using the Jaffe method. Estimated GFR was calculated using the race-free 2021 CKD-Epidemiology Collaboration (CKD-EPI) equation if participant was over age 20 (24), and the Schwartz-Lyon equation if the participant was aged 15 to 20 years (25). The Schwartz-Lyon equation was chosen due to its validity in estimating GFR in adolescents, and CKD-EPI based on KDIGO 2023 recommendations (3). We stratified by risk categories (low to very high risk) using eGFR and urine albumin creatinine ratio (UACR) based on KDIGO guidelines. (Supplemental Figure 1). (3)

The urine albumin was measured at the UWI Chemical Pathology Laboratory from the early morning urine specimen with concentration based on the turbidity reaction produced on addition of Sulphosalicylic Acid (SSA). The spot UACR was calculated by dividing the urine albumin concentration by the urine creatinine concentration. Haemoglobin genotyping was performed at the TMRU Laboratory using a plasma sample. These were analysed by alkali haemoglobin electrophoresis on cellulose acetate, and electrophoretic variants were assessed by acid agar gel electrophoresis.

## Inclusion and Exclusion criteria

Inclusion criteria were age ≥5 years, availability of serum creatinine values, urine albumin creatinine ratio, age, sex and height to calculate eGFR. Absence of serum creatinine, sex, height, age or ED for the survey weighting resulted in exclusion from this analysis

The primary outcome was prevalent CKD, defined as reduced eGFR <60ml/min/1.73m^2^ and/or albuminuria (≥30 mg/g).

## Main exposure variables

Hypertension as defined as self-reported hypertension (if the respondent indicated medication use for hypertension) or if mean systolic pressure of three readings ≥140 mmHg or diastolic blood pressure≥ 90 mmHg. Diabetes mellitus was defined as a fasting glucose >7.0 mmol/L or self-reported diabetes (if the respondent indicated medication use for diabetes).

Body mass index (BMI) was calculated and then categorized in World Health Organization weight categories.(14, 20) Sickle cell trait (SCT) and disease were defined using haemoglobin genotyping. Responses to highest education level attained was used to determine education level. Responses were categorized as “less than secondary school” if the response included no education, primary, or junior secondary school (less than grade 10); “Secondary school” if responded with high school education (grades 10-13) and “More than Secondary school” for beyond a high school education. For household socioeconomic status (SES), participants’ responses to ownership of a list of 22 household assets were used. Terciles of the number of household assets were created as follows: Low household SES persons with ≤9 household assets, middle household SES with 10–12 items, and high household SES with 13–22 items. This has been used in prior studies as a determinant of SES in Jamaicans(26).

Smoking status was determined by the participant response to a question on tobacco use, and categorized as “Never Smoked” if response was never smoked, “Former smoker” if response was former smoker and “Current smoker” if response was yes –not every day or yes-daily.

## Sample size and Power

Given the available sample size of 583 participants, we estimated power based on CKD prevalence of 10% (1), and a margin of error of 5%. Given that data analysis accounted for study design, application of a design effect of 1.94 to the sample size of 583 yielded an effective sample size of 301 (21). At the 95% confidence level, this effective sample the study would have 78.3% power to estimate a 10% prevalence with 5% margin of error.

## Statistical Analysis

Data were analysed using STATA software (version 17BE; StataCorp LP).). Sample-based estimates of means and standard deviation for continuous variables, and proportions and frequencies for categorical variables were obtained. Associations for explanatory variables and the outcome of interest using chi-squared tests for proportions and t-test, ANOVA.

Application of sampling weights yielded nationally representative CKD prevalence estimates. The sampling weights were based on the probability of selection of dwellings and EDs, with adjustments for item non-response, and calibration using the Jamaican population distribution in parish- and sex-specific 5-year age bands. Bivariable logistic regression was performed on exposure variables and CKD as the outcome variable. Exposure variables with p-values ≤0.20 on bivariable analyses or a-priori association with CKD were included the multivariable logistic regression model. Hosmer–Lemeshow goodness-of-fit testing was done to assess differences between observed and expected results in the subgroups of the model population.

## Missing Data

A complete case analysis was performed. To determine evidence of the missing at random (MAR) mechanism, we compared the complete case analysis sample data with the remainder of the study sample using two-sample unpaired t-tests and Pearson’s chi-squared tests, as appropriate, in unweighted analyses (Supplemental Table 1) and using Pearson’s chi-squared tests corrected for survey weighting design (Supplemental Table 2).

## Results

### Sample Characteristics

Of 583 participants, 217 (53%) were male. The mean (±SD) age for the sample was 48.5 ± 17.8 years. More than half (58.5%) of the participants were overweight or obese with mean (±SD) BMI of 28.4 ± 9.5 kg/m^2^. Forty-three percent had hypertension, with the mean(±SD) systolic and diastolic blood pressures of 130.6±21.0 mmHg and 83.4±12.1 mmHg respectively. Prevalence of diabetes mellitus was 20.2%, with mean (±SD) fasting glucose of 6.0±2.2mmol/L. Most of the respondents were never-smokers (72.0%). Most had at least high school education (47%), whilst high household socioeconomic status (SES) was noted in one-quarter (25.8%) of respondents. For sex based differences in the sample see table 1.

**Table 1:**
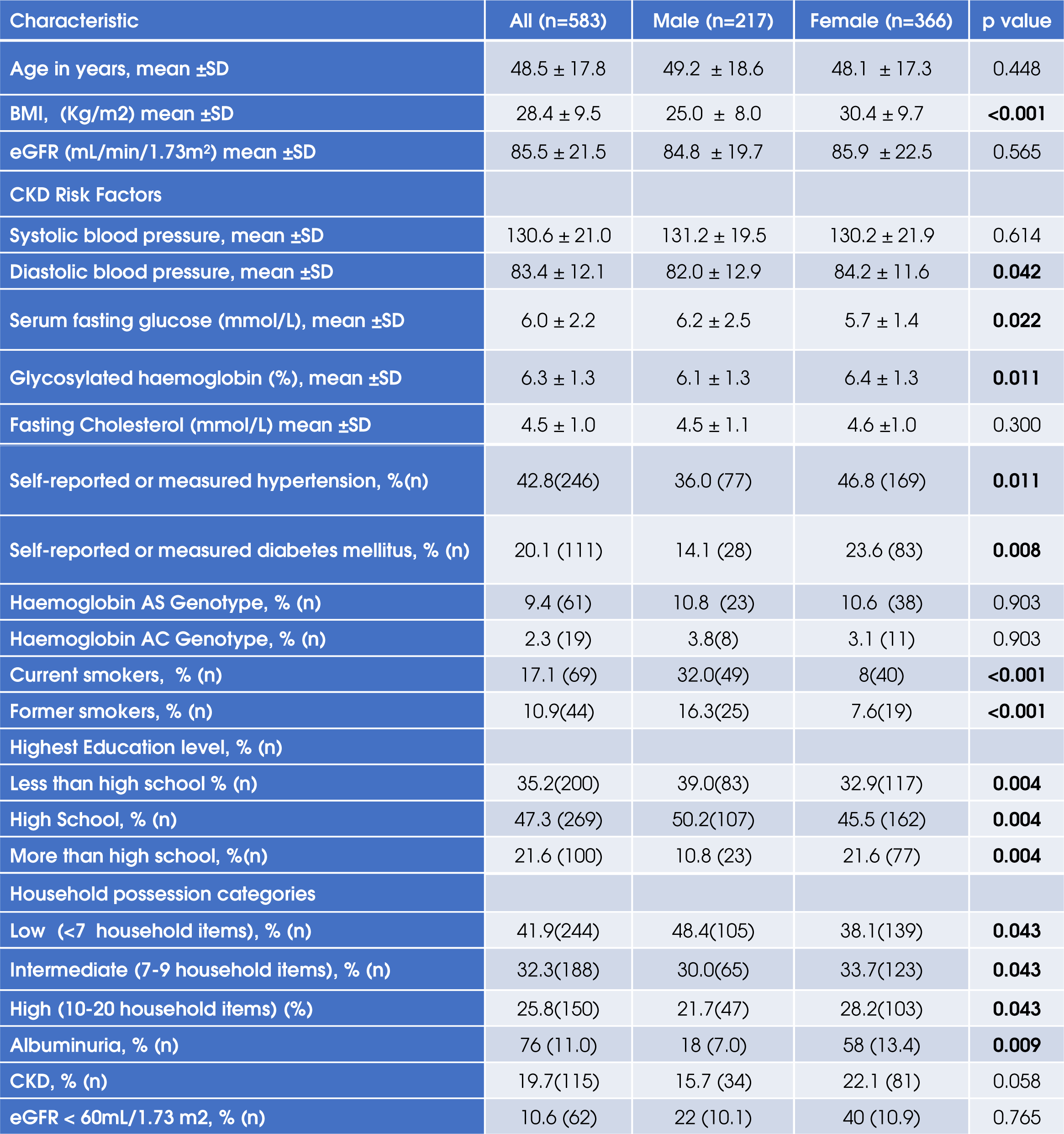
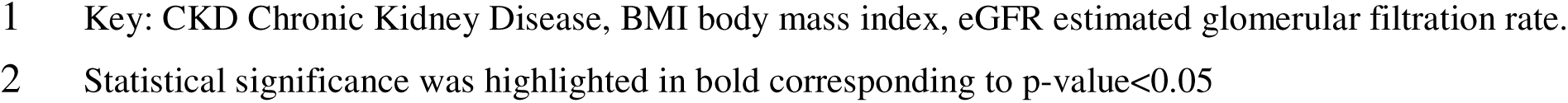
Sex differences in the study population using unweighted estimates.

### Burden of CKD

Based on survey weighting, the prevalence of CKD was 14.8% [95%CI: 11.5%-18.9%]. Just over seven percent (7.2% [95%CI:5.1%-10.1%]) had reduced GFR and 8.8% [95%CI:6.3%-12.0%] had albuminuria. There were no sex differences in survey-weighted CKD prevalence (males: 14.5% versus females 15.0%, p=0.859). Over half (58.5%) of the participants with CKD were >64 years. (Please see Figure 1)

**Figure 1:**
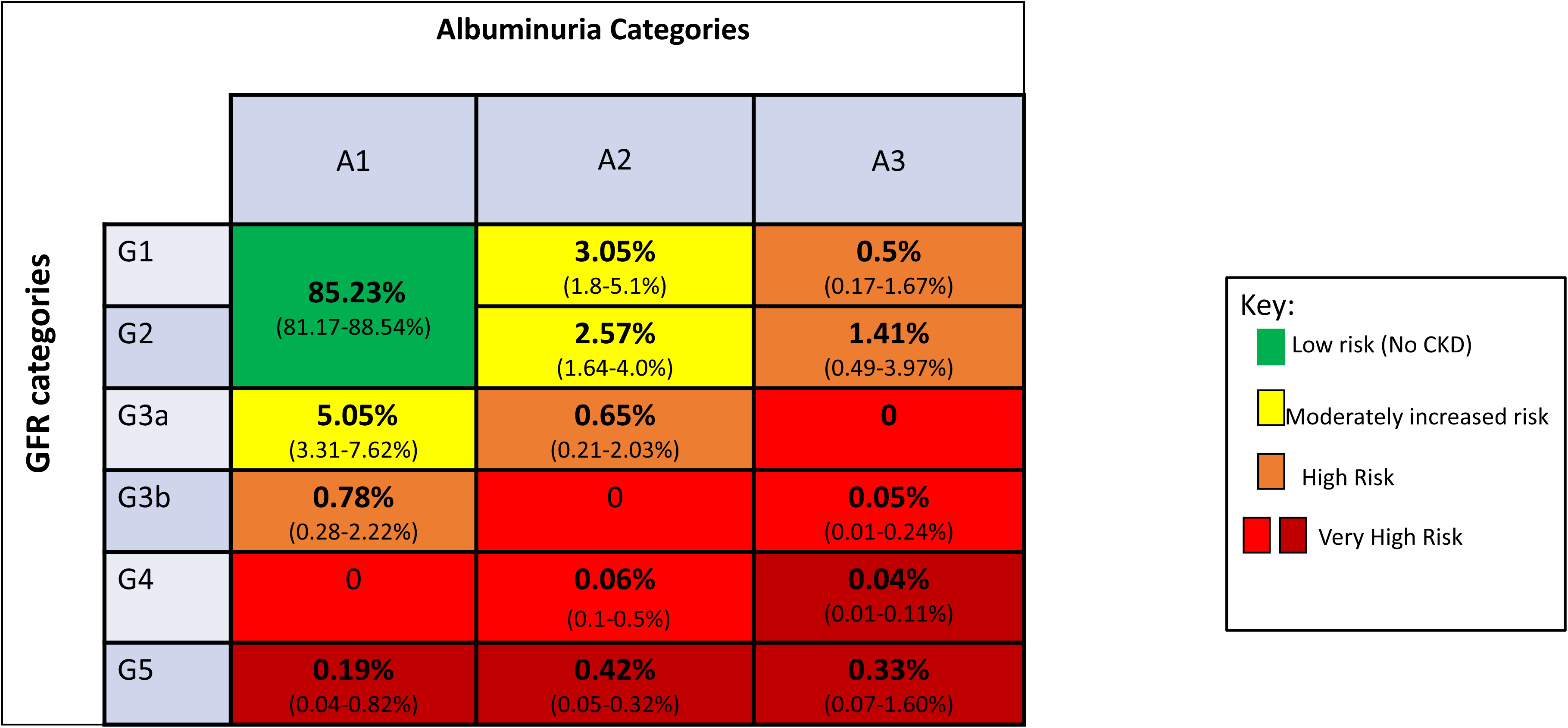
Prevalence of Chronic Kidney disease Stages by Kidney Disease Improving Global Outcomes. Risk Categories. *Bold is percentages based on weighted estimates, with 95% confidence intervals in brackets. CKD= Chronic kidney disease, GFR glomerular filtration rate, A1: urine albumin creatinine ratio (UACR) <30mg/g A2: UACR 30-300mg/g, A3: UACR >300mg/g. G1: GFR* ≧*90mL/min/1.73m^2^, G2: 60-89 mL/min/1.72m^2^, G3a: GFR 45-59mL/min/1.73m^2^, G3b: GFR 30-44mL/min/1.73m^2^, G4 15-29mL/min/1.73m^2^, G5<15 mL/min/1.73m^2^*.

Of those with CKD, the majority had CKD Stage 1 or 2 (46.1%) followed by CKD Stage 3a(40.9%), CKD Stage 3b (7.9%), CKD Stage 4 (1.7%), CKD Stage 5(3.5%). For details of the weighted estimates by KDIGO Stage (see Figure 2). The estimated prevalence of high risk or very high-risk CKD was 4.1% [95%CI: 2.4-6.9].

**Figure 2:**
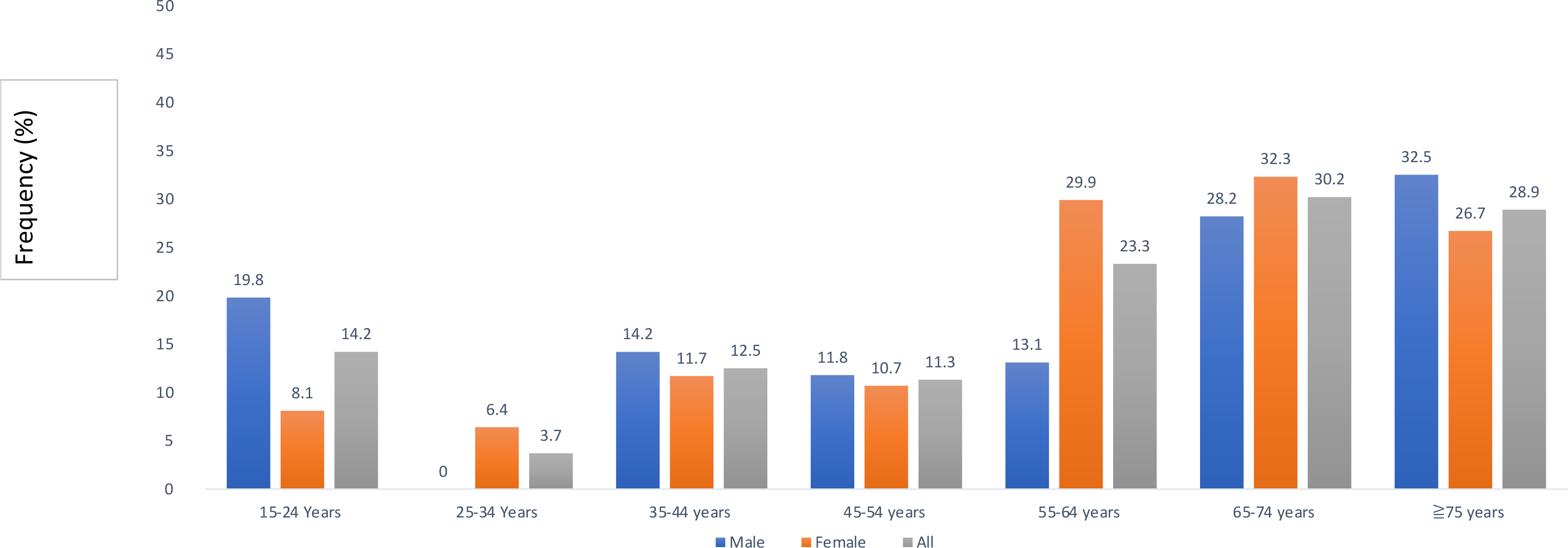
Frequency of Chronic Kidney Disease by 10-year Age Categories.

### CKD Risk Factors

Compared to participants without CKD, persons with CKD were older (mean age 57.0 versus 46.4 years, p<0.001), had higher mean systolic blood pressure (140.3 versus 128.2 mmHg, p<0.001), higher BMI (30.3 versus 27.9 kg/m^2^, p<0.016), higher fasting glucose (6.7 versus 5.8 micromoles/L, p<0.001) and higher glycosylated hemoglobin (6.6 versus 6.2%, p=0.003). There were no statistically significant differences in diastolic blood pressure (85.2 versus 82.9 mmHg. p=0.076) or fasting total cholesterol (4.7 versus 4.5mmol/L, p=0.067) between CKD and non-CKD groups. (see Table 2 for differences by CKD)

**Table 2:**
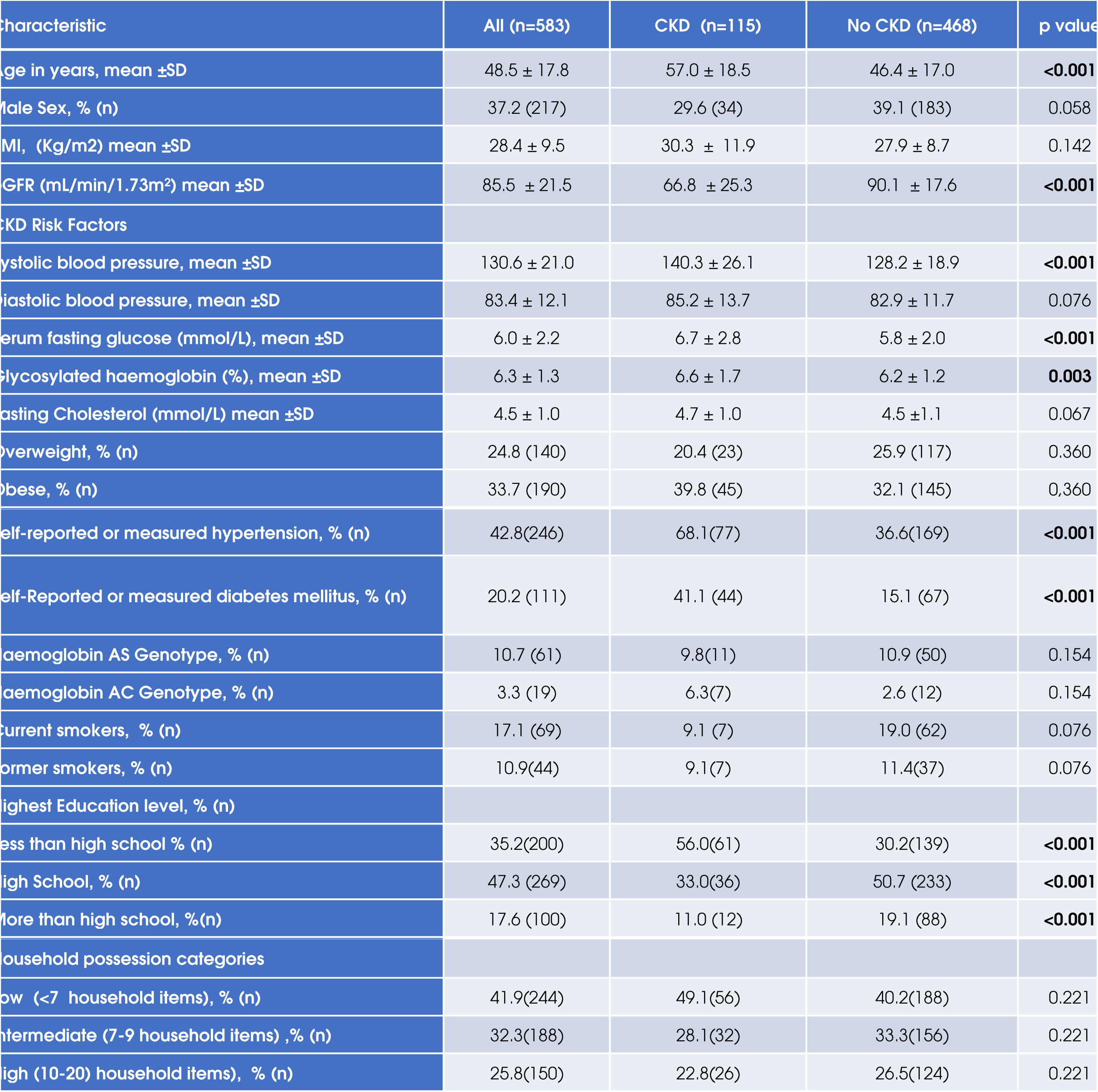
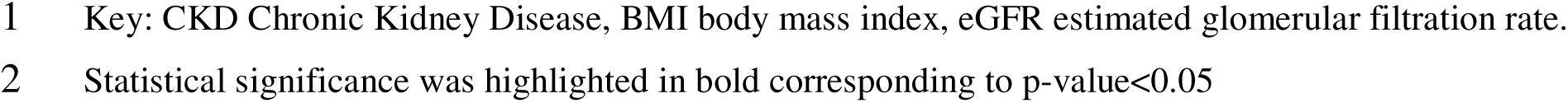
Differences by Chronic Kidney Disease in the Study Population using unweighted estimates.

Self-reported hypertension (68.1% versus 36.5%, p<0.001) and diabetes mellitus prevalence (41.1% versus 15.1%, p<0.001) were higher in participants with CKD. Persons with CKD were less likely to have had secondary school education (56.0% versus 30.2%, p<0.001). There was no statistically significant difference in CKD by sex, household socioeconomic status (SES) or BMI category.

Table 3 presents the final multivariable logistic model. Hypertension (OR 2.14, 95% CI: 1.22-3.76), diabetes mellitus (OR 2.38, 95% CI: 1.36-4.19) were independently associated with CKD. Secondary education(OR 0.47, 95%CI: 0.25-0.89) and tertiary education(OR 0.41, 95CI: 0.18-0.96) were inversely associated with CKD. Hosmer–Lemeshow goodness-of-fit chi-squared statistic for the model was 3.6 (P = 0.731).

**Table 3:** Table with Bivariable and Multivariable logistic regression for Chronic Kidney Disease.

| Variable | Unadjusted Odds Ratio (95% CI) | P value | Multivariable model Odds Ratio (95% CI) | P value |
| --- | --- | --- | --- | --- |
| BMI | 1.02 (1.00-1.04) | 0.020 | 1.01 (0.99-1.04) | 0.333 |
| Age (per 1 year) | 1.04 (1.02-1.05) | <0.001 | 1.01(0.99-1.03) | 0.249 |
| Male sex | 0.65 (0.42-1.02) | 0.059 | 0.79(0.46-1.35) | 0.383 |
| Hypertension | 3.70 (2.39-5.75) | <0.001 | **2.14(1.22-3.76)** | **0.008** |
| Diabetes mellitus | 3.91 (2.46- 6.28) | 0.001 | **2.38 (1.36-4.19)** | **0.003** |
| **Haemoglobin Genotype (reference: Hb AA genotype)** |  |  |  |  |
| Genotype HbAS (Sickle Trait) | 0.93 (0.47-1.85) | 0.359 | 2.14 (0.65-7.13) | 0.215 |
| Genotype Hb AC (HbC carrier) | 2.46 (0.94-6.43) | 0.065 | 1.05 (0.46-2.36) | 0.911 |
| **Education level: (reference: Less than high school)** |  |  |  |  |
| High School | 0.35 (0.22-0.56) | <0.001 | **0.47 (0.25-0.88)** | **0.019** |
| Post High School | 0.31 (0.16-0.61) | 0.001 | **0.41 (0.18-0.96)** | **0.040** |
| **Household possession category: (reference<7 items)** |  |  |  |  |
| 7-9 items | 0.69 (0.43-1.12) | 0.130 | 1.11 (0.61-2.03) | 0.733 |
| 10-20 items | 0.70 (0.42-1.18) | 0.184 | 1.10 (0.56-2.16) | 0.777 |

Key: CI: Confidence Interval, BMI body mass index. Highlighted in bold are the statistically significant $p < 0.05$ on the multivariable logistic regression

### Missing Data

Tables summarizing the characteristics of the sample with missing data on albuminuria and creatinine testing are available in Supplementary Tables 1 and 2. Persons with non-missing data were older, (mean age 48.5 ± 17.8 versus 46.1 ± 18.9 years, p) had higher mean glycosylated haemoglobin (6.3% versus 5.9%, p<0.001), mean fasting glucose (6.0 ± 2.2 versus 5.8 ±2.1, p=0.015), were more likely to have a high household SES (31.1% versus 25.8%, p=0.040) and with a lower proportion who were current smokers (11.6 % versus 17.1%). There were no differences by sex, proportion with diabetes mellitus, hypertension, or obese or overweight, or education level, or haemoglobin genotype. Comparable results were seen in the weighted differences between missing and non-missing groups. A higher proportion persons included in the analysis were in the 45-54- and 55-64-year age groups (18 versus 14.4% and 13.3 versus 6.9% respectively, p=0.03). A higher proportion were obese (32.7 versus 27.3%, p=0.053).

## Data Availability

All data produced in the present work are available upon reasonable request to the authors

## Discussion

In this cross-sectional analysis of Jamaicans, estimated prevalence of CKD, albuminuria and reduced GFR was 15%, 9% and 7 %, respectively. Diabetes and hypertension increased risk whilst higher education attainment reduced risk of CKD.

Our data suggest a higher rate of CKD than prior estimates from GBD and GKHA(1, 11), and is comparable with CKD rates amongst adults in the United States, Canada and Haiti of 13-15%(27-29). Compared to Non-Hispanic Black Americans (19.5%), CKD prevalence was lower(29). Seven percent had impaired kidney function, a rate higher than prior reports from other Caribbean islands of 3 and 4% from Haiti and St Kitts and Nevis respectively(27, 30), but similar to high income countries (HIC) (29). Although these comparisons are less direct because of differences in study methodology (some using survey weighting or age standardized prevalence rates versus crude estimates) and differences in population characteristics(27–30). Despite this, the observed rates of high risk CKD has public health care implications in middle income countries like Jamaica, in which treatment of advanced kidney disease (such as dialysis) are expensive and largely paid out of pocket, resulting in lack of access to care. Jamaica has one of the highest out of pocket payments (51-75%) for dialysis in the Caribbean(10, 13), and a high attributable CKD morbidity and mortality(12).

The high prevalence of CKD, especially high risk CKD in this study, suggest the need for early identification in healthcare systems in Jamaica. Initiation of renin-angiotensin-aldosterone system inhibitors (RAASi), sodium 2 glucose cotransporter inhibitors (SGLT2i), and non-steroidal mineralocorticoid receptor antagonists reduce progression to kidney failure and cardiovascular mortality in CKD(31–33). Addition of SGLT2i to standard of care resulted in a projected 1.7 year longer time with eGFR >15ml/min/1.73m^2^, 1.7 year longer life expectancy with cost effective ratios of between $8280 and $17,623 per quality adjusted life years in 3 European countries(34). Data from South East Asia, and the US also show cost effectiveness and reductions in health care costs and burden with use of SGLT2i and RAASi therapy(35, 36). Focus on screening, at risk individuals, with initiation of early disease modifying therapy may be needed to reduce CKD attributable health care burden. SGLT2i and non-steroidal MRAs are available in Jamaica, but medication costs are prohibitive. CKD is not covered as one of the diseases in the National Health Fund (which subsidizes medications for chronic medical illnesses). This may argue the need for inclusion to improve medication access (10, 13).

Hypertension and diabetes were associated with a twofold risk of CKD, however the higher than expected rates of advanced kidney disease may suggest environmental, genetic or socio-economic factors which affect progression. The presence of high risk Apolipoprotein 1 (*APOL1*) genotypes in persons of African ancestry has been associated with incident CKD and its progression(37–40). Homozygosity for *APOL1* is associated with a 7 to 11 fold increased risk of hypertension attributable ESKF amongst African Americans(39). *APOL1* is associated with lupus nephritis, HIV associated nephropathy and sickle cell related kidney disease(39–42). And is a factor enhancing progression in diabetic kidney disease(39). Jamaica has a predominant West African Ancestry, but no data on *APOL1* high risk prevalence(43), which may warrant further study. Environmental factors both act as a second hit in *APOL1* risk alleles, or are independently associated with CKD(39,43). Ambient temperature, air pollution (particulate matter 25), environmental toxins (e.g. heavy metals), viral infections such as (HIV and dengue virus) are such potential mediators and their role in CKD progression in Jamaica and/or the Caribbean could be examined in future longitudinal cohort studies(43–45). Surprisingly despite a high rate of SCT (11%), there was no association with CKD on multivariable analysis. Data from five population based cohort studies showed a 50% increased risk of incident CKD, and 75% risk of eGFR decline in SCT versus normal genotype, after adjusting for traditional CKD risk factors (46). SCT also increased risk of CKD in persons of African descent residing in the UK with well controlled HIV infection (41). Our lack of association could be explored in further cohort studies.

Higher education attainment, a measure of individual SES, was protective for CKD, congruent with prior observational data showing an inverse association of incident CKD and education(47). Although prior reports have attributed lower prevalence ratios of hypertension and diabetes in Jamaica compared to a US cohort due to differences in age, sex, and BMI rather than education attainment, this difference may be because Jamaican cohort had lower BMIs and had lower rates of hypertension and diabetes and were younger(48). More recent analyses, showed an inverse association of education attainment and multimorbidity independent of other co-variates(49). Education attainment was also associated with five ideal cardiovascular health characteristics (e.g. physical activity, healthy diet and lower BMI) amongst Jamaican women; these factors also reduce CKD rates(21). We did not explore sex-differences in CKD associations, however. That SES was independent of traditional risk factors for CKD, may reflect access to care, health literacy and control of underlying medical disease.

This is the first study estimating CKD prevalence in a large representative sample in Jamaica, however there are several limitations. There was the possibility of selection bias limiting generalizability of results. Representativeness may be compromised by attrition from the primary sample due to missing data. We used creatinine to estimate GFR, the former can be affected by non-GFR determinants of creatinine (e.g. high protein diet, creatine supplements), whilst cystatin-C improves the accuracy of eGFR in persons of African descent(3, 49, 50).

CKD prevalence in Jamaica is comparable to worldwide estimates, but with a high proportion with high risk CKD. Accessible systems for renal replacement therapy coupled with public health measures to enhance early detection and management of CKD are needed.

### Data availability statement

Data for this manuscript is available on reasonable request.

## Acknowledgements

We would like to thank the Ministry of Health and Wellness and the National Health Fund Jamaica, which provided funding for the Jamaica Health and Lifestyle Survey III in 2016/17.We would also like to thank the JHLS-III field staff, JHLS-III participants, CAIHR/ERU administrative staff, for their support. We acknowledge the work of the Jamaica Health and Lifestyle Collaborators who conceived and contributed to the original study. JHLS-III Collaborators (listed alphabetically) Christopher Charles, Sharmaine Edwards, Nicholas Elias, Kelly-Ann Gordon-Johnson, Georgiana Gordon-Strachan, Ardene Harris, Jennifer Knight-Madden, Tiffany Palmer, Suzanne Soares-Wynter, Katherine Theall, Iyanna Wellington, Jovian Wiggan, Janeil Williams, Shara Williams-Lue

## Authors contributions

LAF wrote the first draft of the manuscript and performed the final analysis. RJW, TSF, MTR conceived the original study. SM, TD, CCM, DF, WA, NYC, DM, NGD, KR collected the data and interpreted the results. TSF, NGD, MTR, SM, NYC, KR supervised the analysis. All Authors revised and approved the final version of the manuscript.

