## Supplemental Table 1 for "Chronic Kidney Disease in Jamaica: Estimated Prevalence and Associated Risk Factors from the Jamaica Health and Lifestyle Survey III"

Supplemental Table 1: Table highlighting differences between Chronic Kidney Disease subset and the Jamaica Health and Lifestyle III subset with missing data

| **Characteristic** | **JHLSIII sample (n=2807)** | **Non Missing Data (n=583)** | **Missing Data (n=2224)** | **p value** |
| --- | --- | --- | --- | --- |
| **Age in years, mean ±SD** | 46.0 ± 19.2 | 48.5 ± 17.8 | 46.5 ± 18.9 | **0.006** |
| **BMI, (Kg/m2) mean ±SD** | 27.9 ± 8.5 | 28.4 ± 9.5 | 27.8 ± 8.2 | 0.142 |
| **Male Sex, % (n)** | 38.9 (1093) | 37.2 (217) | 39.3(876)) | 0.355 |
| **CKD Risk Factors** |  |  |  |  |
| **Systolic blood pressure, mean ±SD** | 131.5 ± 21.9 | 130.6 ± 21.0 | 131.8 ± 22.1 | 0.244 |
| **Diastolic blood pressure, mean ±SD** | 83.6 ± 12.4 | 83.6 ± 12.4 | 82.9 ± 11.7 | 0.076 |
| **Serum fasting glucose (mmol/L), mean ±SD** | 5.8 ± 2.1 | 6.0 ± 2.2 | 5.8 ± 2.1 | **0.015** |
| **Glycosylated haemoglobin (%), mean ±SD** | 6.0 ± 1.3 | 6.3 ± 1.3 | 5.9 ± 1.3 | **<0.001** |
| **Fasting Cholesterol (mmol/L) mean ±SD** | 4.5 ± 1.0 | 4.5 ± 1.0 | 4.5 ±1.0 | 0.910 |
| **Overweight, % (n)** | 35.6 (624) | 24.8 (140) | 25.6 (484) | 0.821 |
| **Obese, % (n)** | 32.4 (795) | 33.7 (190) | 32.0 (25.6) | 0.821 |
| **Self-reported or measured hypertension, % (n)** | 44.9 (1215) | 42.8(246) | 45.5 (969) | 0.254 |
| **Self-Reported or measured diabetes mellitus, % (n)** | 18.6 (400) | 20.2 (111) | 18.0 (289) | 0.265 |
| **Haemoglobin AS Genotype, % (n)** | 11.2(137) | 10.7 ( 61) | 11.5 (80) | 0.754 |
| **Haemoglobin AC Genotype, % (n)** | 3.0 (37) | 3.3 (19) | 18 (2.7) | 0.754 |
| **Current smokers, % (n)** | 12.8 (238) | 17.1 (69) | 11.6 (169) | **0.013** |
| **Former smokers, % (n)** | 11.7 (217) | 10.9(44) | 11.9 (173) | **0.013** |
| **Highest Education level, % (n)** |  |  |  |  |
| **Less than high school % (n)** | 33.0 (893) | 35.2(200) | 32.4 (693) | 0.330 |
| **High School, % (n)** | 47.6 (1288) | 47.3 (269) | 47.7 (1019) | 0.330 |
| **More than high school, %(n)** | 19.4 (524) | 17.6(100) | 19.9 (424) | 0.330 |
| **Household possession categories** |  |  |  |  |
| **Low (<7 household items), (%)** | 40.3 (1125) | 41.9(244) | 39.8 (881) | **0.040** |
| **Intermediate (7-9 household items) (%)** | 29.8 (832) | 32.3(188) | 29.1(644) | **0.040** |
| **High (10-20 household items) (%)** | 30.0 (838) | 25.8(150) | 31.1 (688) | **0.040** |
