## Supplemental Table 2 for "Chronic Kidney Disease in Jamaica: Estimated Prevalence and Associated Risk Factors from the Jamaica Health and Lifestyle Survey III"

Supplemental Table 2: Table highlighting differences between Chronic Kidney Disease subset and the Jamaica Health and Lifestyle III subset using with missing data using survey weighting

| **Characteristic** | **JHLSIII sample (n=2807)** | **Non-Missing Data (n=583)** | **Missing Data (n=2224)** | **p value** |
| --- | --- | --- | --- | --- |
| **Age Categories** | | | | |
| **15-24 years, %** | 25.5 | 22.3 | 26.3 | **0.030** |
| **25-34 years, %** | 21.0 | 17.8 | 21.9 | **0.030** |
| **35-44 years, %** | 17.3 | 16.4 | 17.6 | **0.030** |
| **45-54 years, %** | 15.2 | 18.0 | 14.4 | **0.030** |
| **55-64 years, %** | 10.0 | 13.3 | 9.1 | **0.030** |
| **65-74 years, %** | 6.2 | 6.9 | 5.9 | **0.030** |
| **>75 years, %** | 4.9 | 5.4 | 4.8 | **0.030** |
| **Male sex, %** | 48.7 | 45.1 | 49.7 | 0.145 |
| **CKD Risk Factors** | | | | |
| **Overweight, %** | 25.2 | 26.8 | 24.7 | **0.053** |
| **Obese, %** | 28.6 | 32.7 | 27.3 | **0.053** |
| **Self-reported or measured hypertension, %** | 33.8 | 33.7 | 33.8 | 0.952 |
| **Sefl-Reported or measured diabetes mellitus, %** | 13.0 | 13.9 | 12.6 | 0.468 |
| **Hemoglobin AS Genotype, %** | 11.1 | 10.6 | 11.5 | 0.905 |
| **Hemoglobin AC Genotype, %** | 2,5 | 2.6 | 2.5 | 0.905 |
| **Current smokers, %** | 14.6 | 17.9 | 12.4 | 0.141 |
| **Former smokers, %** | 9.9 | 9.8 | 9.8 | 0.141 |
| **Highest Education level** | | | | |
| **Less than high school %** | 22.9 | 23.9 | 22.6 | 0.748 |
| **High School, %** | 53.7 | 54.0 | 53.6 | 0.748 |
| **More than high school, %** | 23.4 | 22.1 | 23.8 | 0.748 |
| **Household possession categories** | | | | |
| **Low (<7 household items),** | 33.0 | 35.7 | 32.2 | 0.061 |
| **Intermediate (7-9 household items) %** | 31.5 | 34.9 | 30.5 | 0.061 |
| **High (10-20 household items) ,%** | 35.6 | 29.5 | 37.3 | 0.061 |

Key: BMI body mass index, CKD chronic kidney disease, JHLSIII Jamaica Health and Lifestyle III
