## Supplemental Figure 1 for "Chronic Kidney Disease in Jamaica: Estimated Prevalence and Associated Risk Factors from the Jamaica Health and Lifestyle Survey III"

### Slide 1
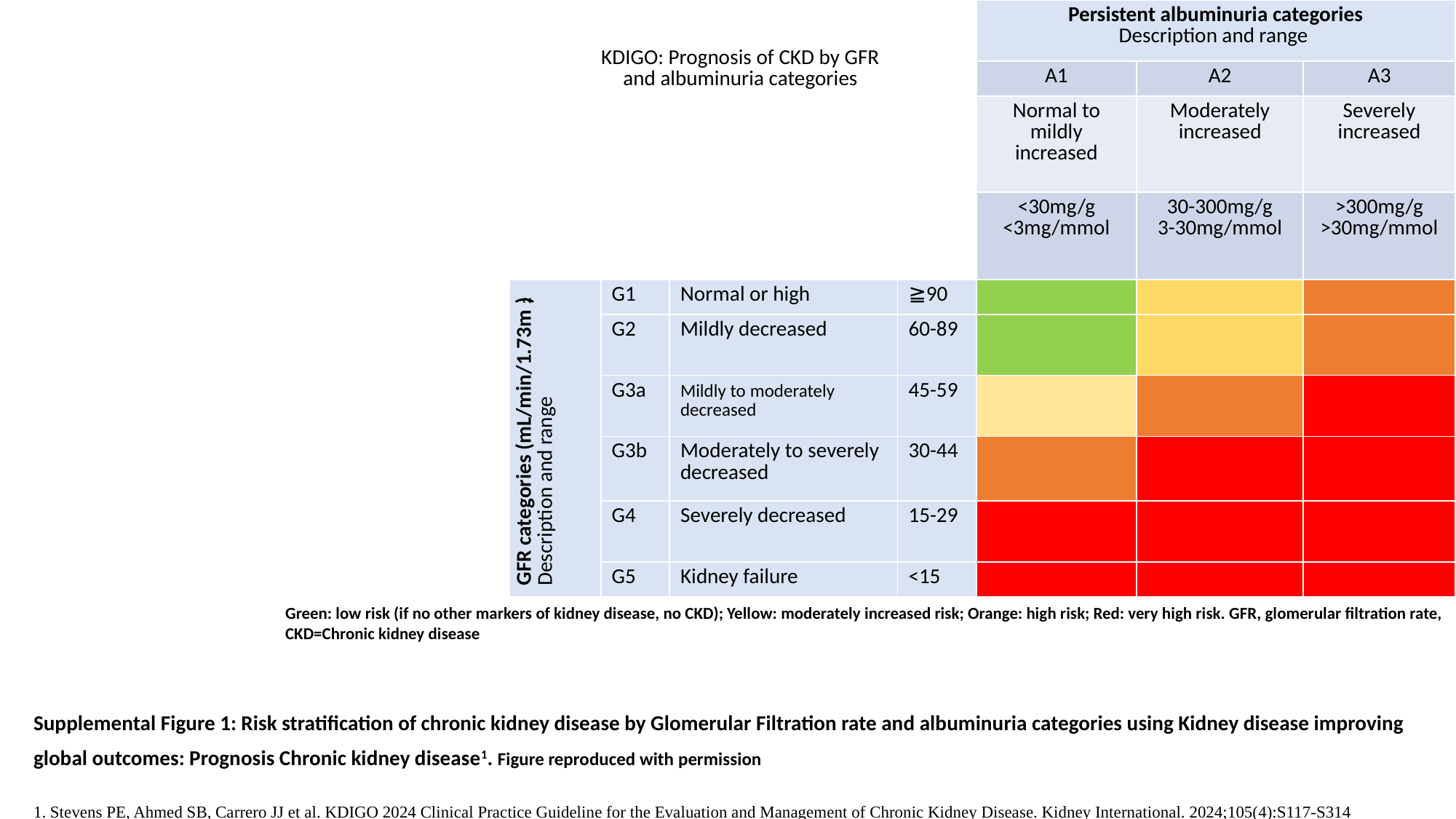

| KDIGO: Prognosis of CKD by GFR and albuminuria categories | | | | Persistent albuminuria categories Description and range | | |
| --- | --- | --- | --- | --- | --- | --- |
| | | | | A1 | A2 | A3 |
| | | | | Normal to mildly increased | Moderately increased | Severely increased |
| | | | | <30mg/g <3mg/mmol | 30-300mg/g 3-30mg/mmol | >300mg/g >30mg/mmol |
| GFR categories (mL/min/1.73m^2^) Description and range | G1 | Normal or high | ≧90 | | | |
| | G2 | Mildly decreased | 60-89 | | | |
| | G3a | Mildly to moderately decreased | 45-59 | | | |
| | G3b | Moderately to severely decreased | 30-44 | | | |
| | G4 | Severely decreased | 15-29 | | | |
| | G5 | Kidney failure | <15 | | | |
Green: low risk (if no other markers of kidney disease, no CKD); Yellow: moderately increased risk; Orange: high risk; Red: very high risk. GFR, glomerular filtration rate, CKD=Chronic kidney disease
Supplemental Figure 1: Risk stratification of chronic kidney disease by Glomerular Filtration rate and albuminuria categories using Kidney disease improving global outcomes: Prognosis Chronic kidney disease^1^. Figure reproduced with permission
1. Stevens PE, Ahmed SB, Carrero JJ et al. KDIGO 2024 Clinical Practice Guideline for the Evaluation and Management of Chronic Kidney Disease. Kidney International. 2024;105(4):S117-S314
